# CADA-PRO, a patient questionnaire measuring key cognitive, motor, emotional and behavioral Outcomes in CADASIL

**DOI:** 10.1101/2024.05.30.24306664

**Authors:** Cécile Di Folco, Aude Jabouley, Sonia Reyes, Carla Machado, Stéphanie Guey, Dominique Hervé, Fanny Fernandes, Joseph Agossa, Hugues Chabriat, Sophie Tezenas du Montcel

## Abstract

**Background:** Cerebral Small Vessel Disease (cSVD) of ischemic type, either sporadic or genetic, as Cerebral Autosomal Dominant Arteriopathy with Subcortical Infarcts and Leukoencephalopathy (CADASIL), can impact the quality of daily life on various cognitive, motor, emotional or behavioral aspects. No instrument has been developed to measure these outcomes from the patient’s perspective. We thus aimed to develop and validate a patient-reported questionnaire.

**Methods:** In a development study, 79 items were generated by consensus between patients, family representatives and cSVD experts. A first sample of patients allowed assessing the feasibility (missing data, floor and ceiling effect, acceptability), internal consistency, and dimensionality of a first set of items. Thereafter, in a validation study, we tested a reduced version of the item set in a larger sample to assess the feasibility, internal consistency, dimensionality, test-retest reliability, concurrent validity, and sensitivity to change.

**Results:** The scale was developed in 44 cSVD patients and validated in a second sample of 89 individuals (including 43 patients with CADASIL and 46 with another cSVD). The final CADASIL Patient-Reported Outcome (CADA-PRO) scale comprised 18 items covering four categories of consequences (depression/anxiety, attention/executive functions, motor, daily activities) of the disease. The proportion of missing data was low, no item displayed major floor or ceiling effect. Both the internal consistency and test-retest reliability were good (Cronbach alpha = 0.95, intraclass correlation coefficient = 0.88). In patients with CADASIL, CADA-PRO scores correlated with the modified Rankin scale, Starkstein Apathy Scale (SAS), Hospital Anxiety and Depression scale (HAD), and Trail Making Test times. In patients with other cSVDs, CADA-PRO correlated only with HAD and SAS.

**Conclusion:** The CADA-PRO is an innovative instrument for measuring patient-reported outcomes in future cSVD trials. Full validation was obtained for its use in CADASIL patients, but further improvement is needed for its application in other cSVDs.

## Introduction

Cerebral small vessel disease (cSVD) is a leading cause of stroke, cognitive decline and disability developing with aging (1). Alongside sporadic forms of cSVD, whose prevalence in the general population is considerable (1), various genetic cSVDs have been identified since the 1990s (2). Among these, Cerebral Autosomal Dominant Arteriopathy with Subcortical Infarcts and Leukoencephalopathy (CADASIL) is, by far, the most frequently diagnosed hereditary cSVD worldwide (3). The disease is caused by typical cysteine mutations of the NOTCH3 gene which encodes a receptor, expressed only at the surface of smooth muscle cells or pericytes, in the wall of arterioles and capillaries (3–7).

CADASIL is now recognized as a paradigmatic cSVD of ischemic type (3). The disease recapitulates most of the clinical and imaging manifestations observed in multiple sporadic or genetic forms of cSVDs. During the past 30 years, important efforts have been devoted to unravelling the intimate mechanisms of the vascular changes behind the age-related accumulation of tissue lesions related to disease worsening (3,4,8,9). In preclinical CADASIL mouse models, key disease mechanisms have been now deciphered. The accumulation of extracellular domains of NOTCH3 (Notch3ECD) in the vascular wall, involving both the mutant and wild-type NOTCH3 receptor, has been recently found to be the major driver of the segmental loss of smooth muscle cells developing with aging in the cerebral microvasculature (10). Various therapeutic approaches to reduce this accumulation can now be envisaged to obtain disease-modifying treatments in the next future.

In clinical terms, however, a number of obstacles still need to be overcome to assess future therapies in CADASIL. Although the condition is considered as an archetypal cSVD, CADASIL remains a rare disorder (11). It evolves variably and over multiple decades. The clinical spectrum is broad and may include acute or chronic symptoms, sensorimotor, cognitive, mood or even behavior manifestations (3,12). These difficulties are further complicated by the varying degrees of symptom awareness felt by the patients themselves, apprehension depending on the family’s experience of the disease, or support by relatives or socio-medical resources. In the presence of such protean clinical manifestations and complex personal, family and social consequences, not only the right treatment should be tested for the right person, at the right time, but it will be just as crucial to show in a robust manner that the treatment addresses clinically relevant issues that are actually meaningful to the patients (13). Patient Reported Outcomes Measures (PROM) are increasingly developed exactly for this purpose and become recommended to support claims in approved medical product by regulatory agencies (14,15).

In the present study, we aimed to assess the first self-reported questionnaire developed to capture different patient reported outcomes in cSVD. The tool that we called CADA-PRO was developed in collaboration with CADASIL patients, family representatives, psychologists, and clinicians to cover multidimensional consequences of the disease on daily living at early or intermediate stage of the disease. The tool properties were investigated according to the COnsensus-based Standards for the selection of health status Measurement INstruments (COSMIN) guideline (16).

## Methods

### General study plan

The feasibility and validity of the CADA-PRO scale were assessed using a two-stage procedure comprising a development study and a validation study. Two distinct samples of patients were used for these two studies.

In the development study, multiple questions or items were first generated by experts, patients and family members. Thereafter, a first sample of patients diagnosed with CADASIL or other cSVD was recruited to assess the feasibility, internal consistency, and dimensionality of this first set of items. Patients’ caregivers were also recruited to evaluate a caregiver version of the set of items.

In the validation study, a reduced set of items was chosen and assessed in a larger sample of patients for the feasibility, internal consistency, reliability, dimensionality, concurrent validity, and sensitivity to change.

The psychometric properties of the final set of items were finally analyzed.

### The CADA-PRO development study

#### Participants

Patients participating to the development study were enrolled from January 2020 to May 2021. They were recruited at day hospital or during outpatient consultations planned for work-up after a recent stroke event or an MRI-based diagnosis of ischemic cSVD (most often sporadic cSVD but also CADASIL). The recruitment target was fixed at 50 patients. Additional inclusion criteria were: 1) Age > 18 years, 2) French native speaking and literacy, 3) Independent ability to complete questionnaires, 4) confirmed diagnosis of a typical ischemic cSVD (white matter hyperintensities with or without lacunes or microbleeds) at MRI examination. Exclusion criteria were non-acquired cognitive disability, nonvascular leukoencephalopathy, suspicion of degenerative disease, severe or unstabilized psychiatric pathologies (psychosis, severe depression), and unstable clinical state (seizures or recent stroke).

To evaluate the patient caregiver’s questionnaire, individuals were selected if they were easily reachable, informative and in contact with the patient at least once every 15 days.

#### Setting up the initial questionnaire

All items of the questionnaire were first developed together by 1) four psychologists with extensive experience in assessing patient complaints, listening to difficulties of patients, caregivers, and families and evaluating cognitive performances and neuropsychiatric disturbances (CM, AJ, MHD, SR), 2) three to four patients (or their representatives) belonging to the CADASIL French Family Association (CADASIL-France) whose aim is to inform and help patients and their families to cope with the disease, 3) two neurologists (HC, DH) having a long experience in patient care and follow-up. These items were chosen to evaluate cognitive functioning, emotional and behavioral symptoms, as well as motor disturbances that can impact activities of daily living. After in-depth discussion during repeated meetings involving all (n = 2) or some members of these groups (n = 4), 79 Likert-scaled items were selected and sorted (Supplement Table 1).

#### Questionnaire administration and evaluation

During the questionnaire administration performed at hospital, study participants were asked whether the selected items were understandable, accurately, and exhaustively represented their everyday difficulties with the disease, could be answered without assistance, and were emotionally difficult to answer. Items that did not meet these acceptability criteria were removed. Individuals were also asked to give out cognitive, motor, emotional, behavioral, and everyday difficulties that would be missing from these first questions.

#### Data Analysis

We first analyzed the characteristics of patients included in the study, the frequency of missing data, and floor and ceiling effects on each item. The internal consistency of the questionnaire was evaluated using Cronbach’s alpha and its structure was assessed using an Exploratory Factor Analysis (EFA). Prior to EFA, missing data were imputed with Multivariate Imputation by Chained Equations (MICE) algorithm.

### The CADA-PRO validation study

#### Participants

Patients participating to the validation study were enrolled from November 2021 to February 2023. They were recruited and selected according to the same procedures as those used in the development study. The initial recruitment target was 100 patients.

#### Setting up the validation questionnaire

The validation questionnaire was prepared based on the results of the development study. Each item of the first questionnaire was kept for the validation study when it fulfilled the following criteria: 1) less than 20% of missing data, 2) less than 60% of answers at floor or ceiling modality, 3) did not decrease the Cronbach’s alpha of the set of items, 4) highest loading among items measuring the same trait.

#### Questionnaire administration and evaluation

All participants completed the CADA-PRO questionnaire at hospital on the day of inclusion in the validation study, one month later when they were alone at home (M1), and one year later during a follow-up visit at hospital (M12).

To establish the external validity of the final questionnaire, an extensive clinical evaluation was performed at the day of inclusion and during the one-year follow-up visit at hospital. This included a large battery of cognitive tests and a global assessment of disability, motor disturbances, mood, and behavior. Order between the CADA-PRO completion and neuropsychological assessments was randomized.

Mental flexibility and processing speed were evaluated using seven scores. One was the number of correct answers at the VADAS-Cog Symbol Digit Test (18–20). Four were obtained from the Trail Making Test (TMT): the time for completion of TMT part A (TMT A time) and TMT part B (TMT B time), their difference (TMT B-A time), and the Number of Errors in TMT part B (21,22). Two were obtained from the Wisconsin Card Sorting Test (WCST) (23): the number of completed categories and the number of perseverations. Working memory was assessed using the Working Memory Index from the Weschler Memory Scale 3rd revision (WMS-III) (24). Verbal memory performance was analyzed using scores obtained from the Free and Cued Selective Reminding Test (FCSRT) adapted from the Grober and Buschke procedure (25): the total Free Recall and total Cued Recall scores (sum of 3 trials each varying from 0 to 16 [best score]), as well as the Index of Sensitivity to Cueing (a measure of retrieval/storage ability that decreases when information storage was compromised and cues are not useful) (26).

Motor symptoms were evaluated using 4 items from the Short Physical Performance Battery (SSPB) (27): Standing Balance Test with 1-side by side feet, 2 -feet in Tandem, 3 - single foot stand, and Single Chair Stand Test. A SSPB-4 score was computed as the sum of these four items (4 – best score, 0 – worst score).

Emotional and behavioral symptoms were evaluated with the Hospital Anxiety and Depression scale (HAD) (28), and the Starkstein Apathy Scale (SAS) (29).

Disability was assessed using the modified Rankin scale. Daily living activities and functional independence were evaluated globally using the Instrumental Activities of Daily Living (IADL) scale (30).

At the final visit, patients were further asked to complete the Patient Global Impression of Change (PGIC) (17).

#### Data analysis

We first analyzed the main characteristics of all participants in the validation study at inclusion, M1 and M12. We also compared these characteristics between patients with CADASIL and patients with other types of cSVD.

As for the development study, we checked the frequency of missing data, floor and ceiling effects on each item. More specifically, the different items were kept if they: 1) had less than 20% of missing data, 2) had less than 60% of answers at floor or ceiling modality, 3) did not decrease the Cronbach’s alpha of the set of items.

To analyze the properties and structure of the validation questionnaire, we used a Confirmatory Multidimensional Item Response Theory Model. Items with insufficient communalities (<0.4) were removed. The item selection process is summarized in Supp Figure 1.

**Figure 1:**
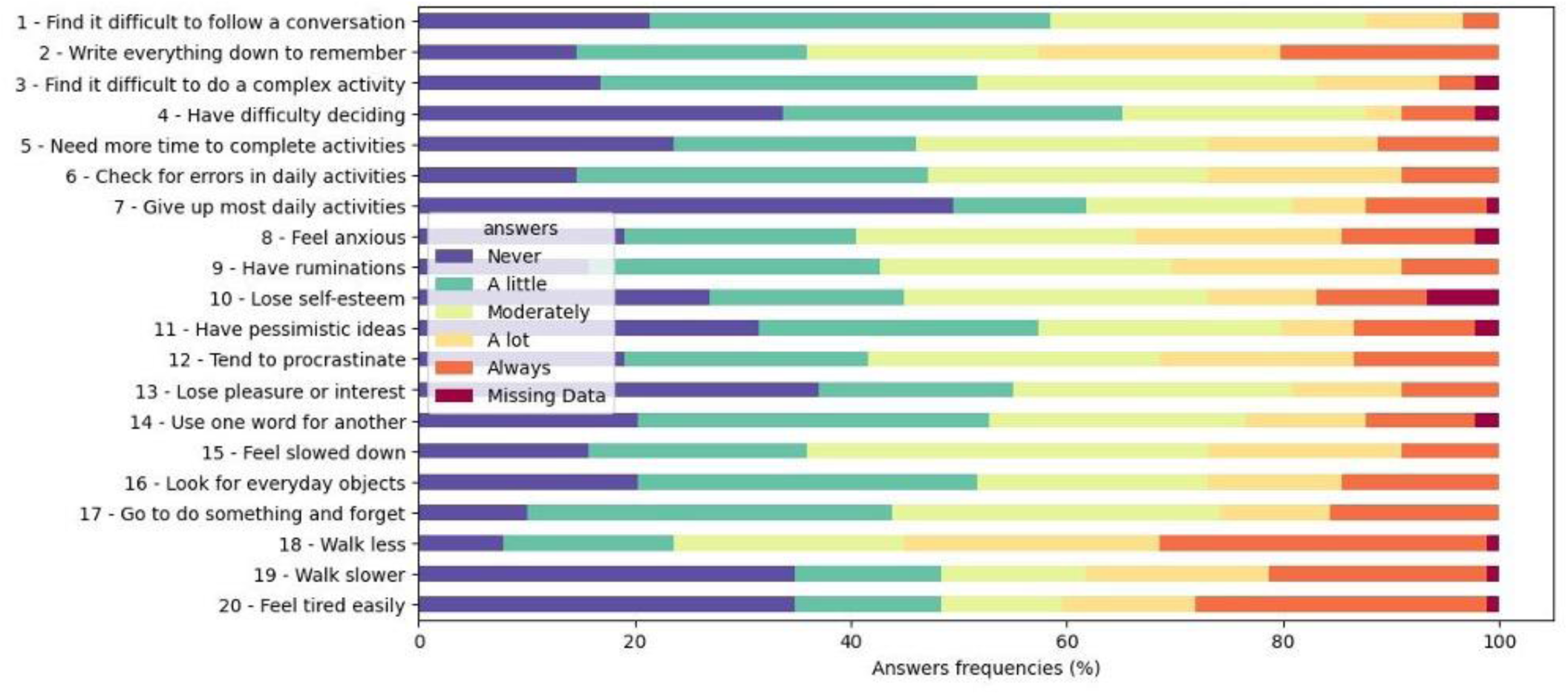
Frequencies of answers obtained at inclusion visit for the selection of items composing the validation-study version of the CADA-PRO tool (N=89)

Then, we computed the total CADA-PRO score as the unweighted sum of all questionnaire items. In addition, for each factor estimated in the structure analysis, we calculated a sub-score as the unweighted sum of the corresponding items. The total score was not computed for patients having more than 20% of missing data. For patients with less than 20% missing data, if only one value was missing per sub-score, it was imputed using the average of the items belonging to the same sub-score. Patients for whom the CADA-PRO total score could not be computed were excluded from the analysis. The effect of order between the CADA-PRO completion and neuropsychological assessments on the CADA-PRO total score was checked.

We also assessed the test-retest reliability between the results obtained at inclusion and at one-month completion using quadratic-weighted Cohen’s Kappas at each item level and the Intraclass Correlation Coefficient (ICC) at the score and sub-score levels. For scores and sub-scores, we further calculated the Standard Error of Measurement (SEM) and Smallest Detectable Change (SDC) between measures obtained at inclusion and one-month completion. SEM was computed as SD × √(1 − ICC), where SD was the standard deviation of all measures of the considered CADA-PRO score. SDC was then computed as SEM × 1.96 × √2 / √n, where n was the number of all measures of the same CADA-PRO score.

We further conducted concurrent validity analysis separately for each diagnosis group (CADASIL or other cSVD). First, we computed the correlations between CADA-PRO scores and clinical scores that had enough variability within our sample (SAS, HAD anxiety and depression, VADAS-Cog code, Working Memory Index from WMS-III, SSPB-4, FCSRT total free recall and reactivity index, TMT B-A). Second, we performed a multiple regression to explain the CADA-PRO total score by the clinical scores selected through a stepwise variable selection.

Finally, we assessed sensitivity to change between inclusion and 1-year completion. We computed the mean score difference between the two assessments, the p-value of a two-tailed T-test testing if this mean score difference was different from 0, as well as the corresponding effect size (mean score difference divided by the standard deviation of score difference).

For two-group comparisons, T-tests were performed on quantitative variables, Mann-Whitney U test on modified Rankin Score, and Chi-square tests on categorical variables. For more than two-group comparisons in quantitative values, ANOVA were performed, and for significant differences, Tukey-Kramer adjusted pairwise comparisons were performed. Data were expressed as mean ± standard deviation or frequency (percent). Statistical tests were performed at the conventional 2-tailed type I error of 0.05. Data were analyzed using R version 4.0.3 (The R Foundation for Statistical Computing, 2020) and Python 3.8.

### Standard Protocol Approval, Registration, and Patient Consents

Informed consent was obtained from each subject or from a close relative if necessary. Data were collected through the SMACS study, that was approved by an independent ethics committee (2019-A01892-55).

### Data Availability

Anonymized data not published within this article will be made available upon request from any qualified investigator.

## Results

### The CADA-PRO development study

Forty-four individuals participated to the CADA-PRO development study using the initial version of the CADA-PRO questionnaire. one patient was diagnosed with CADASIL, 7 had a monogenic cSVD distinct from CADASIL (mutation of COL4A1/COL4A2) and 36 displayed a sporadic form of cSVD related to aging, hypertension or other vascular risk factors. According to their Rankin score, they were 80.5% with no disability, 12.2% with slight disability and 7.3% with a moderate disability. (Table 1).

**Table 1:**
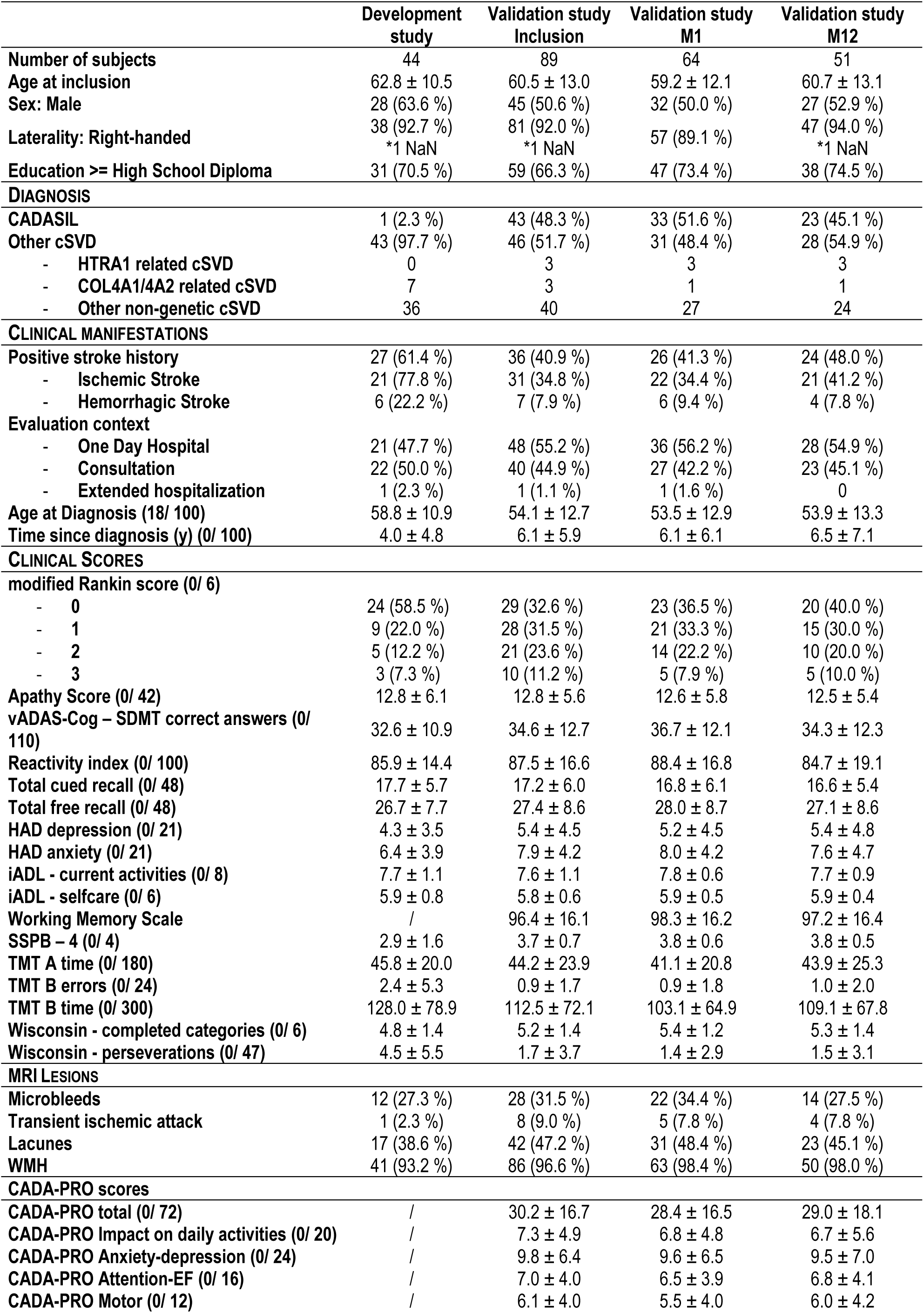
Characteristics at baseline of the participants at each step of the study. **Legend:** The first column shows the characteristics of the participants from the development study. The following columns describe characteristics at inclusion of three sets of participants from the validation study: 1 - those present at inclusion, on which feasibility, internal consistency, dimensionality and concurrent validity were assessed, 2 - those present both at inclusion and M1, on which test-retest reliability was assessed, and 3 - those present both at inclusion and one-year follow-up, used to study sensitivity to change.

Out of the 79 items initially selected for the CADA-PRO questionnaire (Supp Table 1), 11 were excluded due to too many missing data: these items concerned driving and professional activities, which are not relevant for all patients (Supp Figure 2). 26 items were further excluded due to the presence of an obvious floor or ceiling effect (Supp Figure 2). These items mostly assessed severe limitations that are not encountered in the early stage of the disease (for example: need help in daily activities, severe depressive or motor symptoms). Two items were further excluded because they were found to decrease Cronbach’s alpha of the total set of questions. Finally, 20 additional items were excluded due to their insufficient weight in the four first dimensions estimated by the EFA and corresponding to 1) the difficulties in activities of daily living, 2) the changes in attention and executive functions (EF), 3) the development of motor symptoms, and 4) the occurrence of anxiety and depression (Supp Table 3). One item corresponding to a frequent complaint was also added to the final version based on experts’ recommendations (Supp Figure 1).

**Figure 2:**
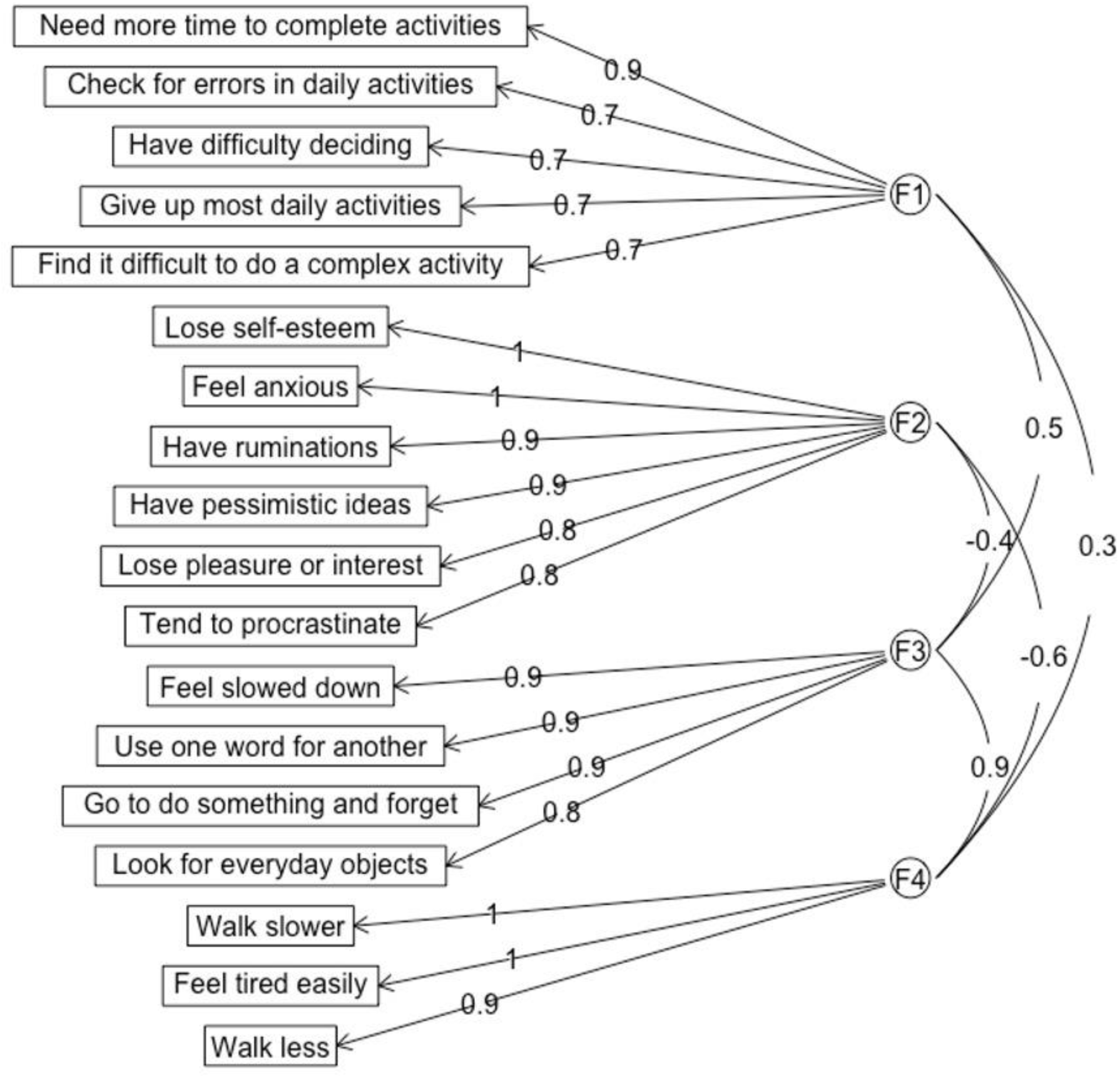
Item loadings and factor correlations for the validation-study version of the CADA-PRO (18-items). <u>Legend</u>: The data are shown after VARIMAX rotation. F1, F2, F3 and F4 are the four identified factors. They are labelled as follows: F1 = Impact on daily activities, F2 = Anxiety/depression, F3 = Attention/EF, F4 = Motor.

As only 70% of patients had a caregiver, and after we observed that caregiver answers were too discordant with those obtained from the patients (Supp Figure 3), the caregiver version of the CADA-PRO questionnaire was not further developed.

**Figure 3:**
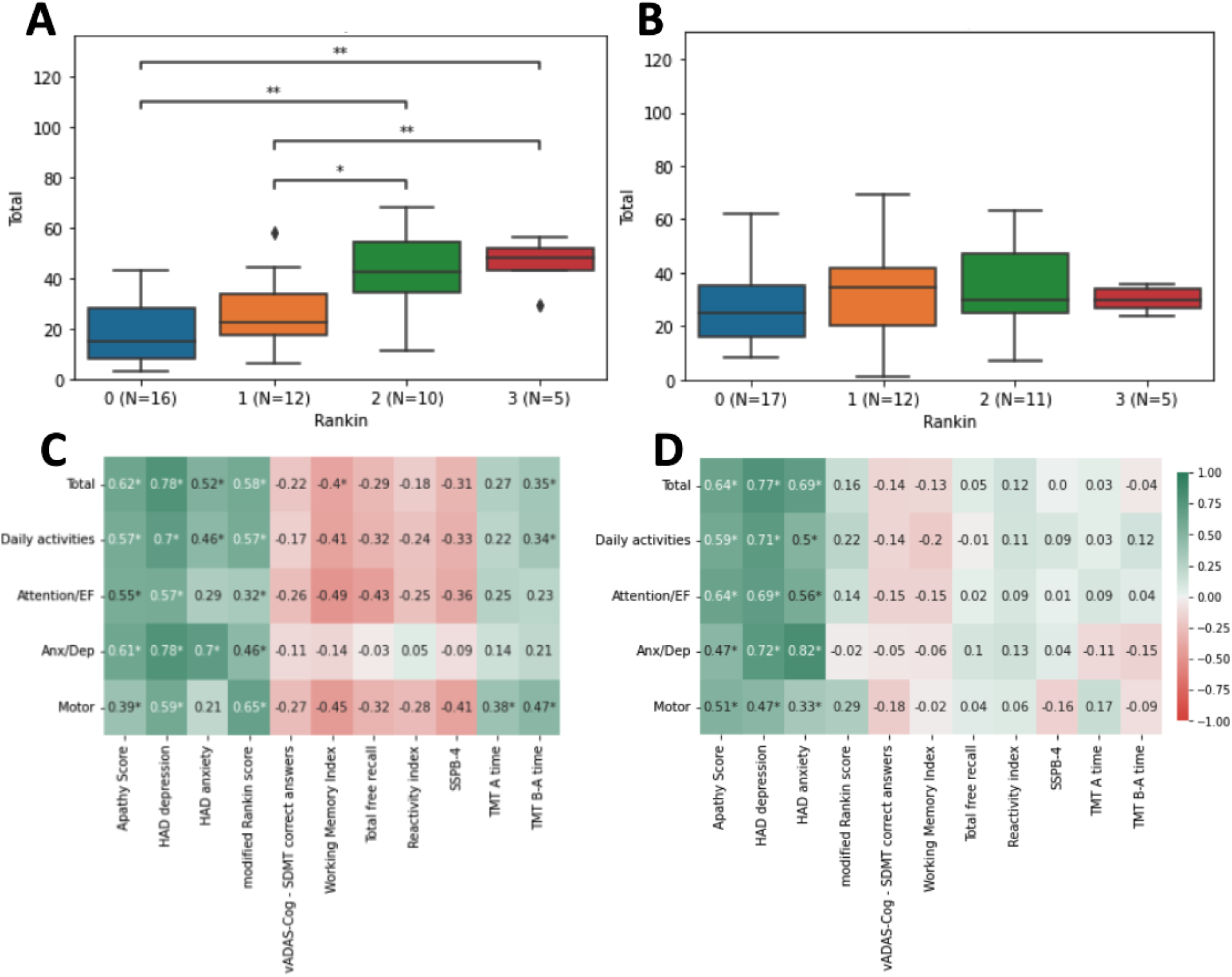
Correlations between the CADA-PRO scores and different clinical scores in patients with CADASIL (A, C) and patients with other types of cSVD (B, D) <u>Legend</u>: A and B: Relationships between that CADA-PRO total score and the Rankin score in CADASIL patients (A) or in patients with another cSVD (B). Significance stars are plotted only in presence of a significant difference between the different levels of the Rankin score. C and D: Pearson correlation coefficients computed between the different CADA-PRO sub-scores and different clinical scores in patients with CADASIL (C) and in those with other cSVDs (D). The CADA-PRO total score is labelled as Total, the Anxiety/Depression sub-score as Anx/Dep, the Impact on Daily Activities sub-score as Daily Activities. Significancy stars are plotted only for significant correlations. Statistical significant is indicated as *: 0.01 < p <= 0.05; **: 0.001 < p <= 0.01; ***: 0.0001 < p <= 0.001; ****: p <= 0.0001.

### The CADA-PRO validation study

Ninety-two patients participated to the CADA-PRO validation study, of whom 89 answered the questionnaire with at least 80% of complete data. Of them, 43 were diagnosed with CADASIL, six had a monogenic cSVD distinct from CADASIL (mutation of COL4A1/COL4A2, N=3; mutation of HTRA1, N=3) and 40 presented with a sporadic form of cSVD related to aging, hypertension or other vascular risk factors. Patients with a cSVD distinct from CADASIL were older at inclusion (56.7 ± 11.8 vs. 64.0 ± 13.1, p=0.007) and at time of diagnosis (49.5 ± 11.7 vs. 59.0 ± 11.9, p<0.001) than CADASIL patients. They also had worse performances for the VADAS-Cog SDMT and Index of sensitivity to cueing from the FCSRT (Supp Table 2). Overall, 63.1% had no disability, 23.6% had a slight disability and 11.2% had a moderate one.

**Table 2:** Internal consistency and test-retest reliability of the final CADA-PRO total score and its four sub-scores. <u>Legend:</u> Internal consistency is assessed by Cronbach’s alpha (N=89). Test-retest reliability (N=64) is assessed by Intraclass correlation coefficient (ICC), Standard Error of Measurement (SEM) and Smallest Detectable Change (SDC).

| **Scores** | **Alpha** | **ICC** | **SEM** | **SDC** |
| --- | --- | --- | --- | --- |
| **Total (0/72)** | 0.95 (0.92, 0.97) | 0.88 (0.80, 0.92) | 5.95 (4.78, 7.56) | 2.06 (1.66, 2.62) |
| **Anx/Dep (0/24)** | 0.91 (0.87, 0.95) | 0.88 (0.80, 0.92) | 2.37 (1.90, 3.01) | 0.82 (0.66, 1.04) |
| **Attention/EF (0/16)** | 0.84 (0.75, 0.90) | 0.69 (0.53, 0.80) | 2.21 (1.77, 2.70) | 0.77 (0.61, 0.94) |
| **Daily Activities (0/20)** | 0.87 (0.80, 0.92) | 0.82 (0.72, 0.89) | 2.09 (1.65, 2.62) | 0.72 (0.57, 0.91) |
| **Motor (0/12)** | 0.85 (0.77, 0.91) | 0.80 (0.70, 0.88) | 1.76 (1.38, 2.19) | 0.62 (0.46, 0.76) |

**Table 3:** Internal consistency, test-retest reliability, and proportion of explained variance for the 18 items of the final CADA-PRO. <u>Legend:</u> Internal consistency (N=89) is assessed by Cronbach’s alpha, for the total set and for the set minus the item. Test-retest reliability (N=64) is assessed by Quadratic-weighted Kappas. The proportion of explained variance by the 4-factor structure (h^2^; N=89) is given, as well as the factor name for which the item was constrained to load onto.

| Item | Cronbach's $\alpha$ | qw-K* | Factor | $h^2$ |
| --- | --- | --- | --- | --- |
| 3 - Find it difficult to do a complex activity | 0.94 (0.92, 0.97) | 0.66 | Daily activities | 0.43 |
| 4 - Have difficulty deciding | 0.94 (0.92, 0.96) | 0.53 | Daily activities | 0.48 |
| 5 - Need more time to complete activities | 0.94 (0.91, 0.96) | 0.75 | Daily activities | 0.84 |
| 6 - Check for errors in daily activities | 0.94 (0.92, 0.96) | 0.66 | Daily activities | 0.55 |
| 7 - Give up most daily activities | 0.94 (0.92, 0.96) | 0.75 | Daily activities | 0.44 |
| 8 - Feel anxious | 0.94 (0.92, 0.96) | 0.71 | Anx/Dep | 0.92 |
| 9 - Have ruminations | 0.94 (0.92, 0.96) | 0.68 | Anx/Dep | 0.87 |
| 10 - Lose self-esteem | 0.94 (0.92, 0.96) | 0.78 | Anx/Dep | 0.93 |
| 11 - Have pessimistic ideas | 0.94 (0.92, 0.96) | 0.76 | Anx/Dep | 0.75 |
| 12 - Tend to procrastinate | 0.94 (0.92, 0.96) | 0.68 | Anx/Dep | 0.63 |
| 13 - Lose pleasure or interest | 0.94 (0.91, 0.96) | 0.79 | Anx/Dep | 0.71 |
| 14 - Use one word for another | 0.94 (0.92, 0.96) | 0.53 | Attention/EF | 0.81 |
| 15 - Feel slowed down | 0.94 (0.91, 0.96) | 0.68 | Attention/EF | 0.86 |
| 16 - Look for everyday objects | 0.94 (0.92, 0.96) | 0.56 | Attention/EF | 0.71 |
| 17 - Go to do something and forget | 0.94 (0.92, 0.96) | 0.60 | Attention/EF | 0.80 |
| 18 - Walk less | 0.94 (0.92, 0.96) | 0.29 | Motor | 0.86 |
| 19 - Walk slower | 0.95 (0.92, 0.97) | 0.78 | Motor | 0.99 |
| 20 - Feel tired easily | 0.95 (0.92, 0.97) | 0.60 | Motor | 0.99 |

\*qw-k = quadratic weighted kappa

#### Feasibility

The administered CADA-PRO questionnaire included 20 items selected from the development study. None of them displayed excessive missing data, floor or ceiling effect (Figure 1). The average proportion of missing data was 1.3% (SD=1.6%, max=6.7% reached on item 10 i.e. Lose self-esteem). The average proportion of answers on the floor modality was 24.1% (SD=10.4%, max = 50.0% reached on item 7 i.e. Give up most daily activities) and that of the ceiling modality was 12.7% (SD=6.9%, max=30% reached on Item 18 i.e. Walk less).

#### Structure

We applied the four-factor structure derived from the development study to the data collected in the validation study. For this purpose, each item was constrained to load on only one latent factor (Supp Table 3). Two items (1– Difficulty following conversation and 2 – Write everything down not to forget) were found to be insufficiently explained by the model (communalities <0.4). They were removed.

The final structure (Figure 2) based on 18 items along 4 main dimensions included: 1) 5 items assessing the impact of the disease on daily activities (“Daily activities”), 2) 6 items related to anxiety and depressive complaints (“Anx/Dep”), 3) 4 items related to impairment in attention and executive functions (“Attention/EF”), and 4) 3 items covering the motor difficulties (“Motor”). These four factors respectively explained 15.2%, 26.7%, 17.7%, and 15.8% of the variance (RMSEA=0.17 (0.15, 0.19), TLI=0.89, CFI=0.91).

The CADA-PRO total score computed as the sum of the 18 different items, ranged from 0 – no complaint to 72 – maximum complaint. The average score was 30.0 (SD=17.2) for patients with CADASIL, and 30.3 (SD=16.3) for patients with other types of cSVDs. The total score did not depend on completion order (neuropsychological testing/questionnaire; data not shown).

#### Internal consistency

The Cronbach’s alpha of final CADA-PRO was 0.95 (0.92, 0.97) (Table 2). No removal of item was increasing the Cronbach’s alpha (Table 3). The Cronbach’s alpha for the different sub-scores ranged from 0.91 (0.87, 0.95) (Depression/Anxiety) to 0.84 (0.75, 0.90) (Attention/EF).

#### Test-retest reliability

The reliability of the total score was high with an ICC between the score obtained at inclusion and at M1 of 0.88 (0.80, 0.92) (p value = 3.5e-22). ICC was above 0.8 for all sub-scores except the Attention/EF one. Fourteen items displayed substantial reliability (kappa > 0.6), three showed moderate reliability (kappa > 0.5), and one showed low reliability (kappa < 0.4) (Table 3). At the group level, the smallest detectable change in the total score was 2.06 points (range: 1.66-2.62).

#### Concurrent validity

The CADA-PRO total score was significantly related to the Rankin score for patients with CADASIL but not for the others (Anova F = 7.03, p-value<0.001; see Figure 3 A and B). For CADASIL patients, the CADA-PRO score significantly correlated with the SAS, HAD anxiety and depression score, and with TMT B-A time. Correlations between the CADA-PRO score and other clinical scores were all higher for CADASIL patients than for patients with other cSVDs, except for HAD-anxiety. For patients with other cSVDs, the CADA-PRO score significantly correlated only with the SAS, HAD-depression, and HAD-anxiety scores.

At the level of the four-domain sub-scores, for patients with CADASIL, the correlations showed different patterns: the Anxiety-Depression score had the highest correlation with HAD anxiety (r = 0.70, p<0.001), depression (r = 0.78, p<0.001) and the SAS scores (r = 0.61, p<0.001), but does not correlate with other scores. In comparison, the Motor score has the highest correlation with the Rankin score (r = 0.65, p<0.001), and tends to correlate with most clinical scores, significantly for TMT A and B-A times (r = 0.38, p = 0.010, and r = 0.47, p = 0.0015, resp.). Attention/EF and Impact on daily activities scores have a correlation pattern similar to that of the Motor score.

To further understand the association between the CADA-PRO total score and the clinical status, we regressed the total score on the different clinical scores using a stepwise selection procedure (Supp Table 4). For patients with CADASIL, we found a significant effect of HAD anxiety (β=4.7 (2.1, 7.4), p=0.001), HAD depression (β=9.6 (6.9, 12.3), p<0.001), and Working Memory Index (β=-2.3 (-4.6, -0.09), p=0.042), accounting for 66.7% of the CADA-PRO score variance. For patients with other cSVDs, only HAD anxiety (β=4.3 (1.7, 6.9), p=0.001) and HAD depression (β=10.5 (7.8, 13.1), p<0.001) significantly explained the CADA-PRO score (R^2^=64%).

#### Sensitivity to change

At the 12-month assessments (N=51), the patients’ CADA-PRO total score had decreased of 1.53 points on average (SD=10.13, p=0.286, ES=-0.15). For patients with CADASIL (N=23), the score had increased of 0.74 points on average (SD=10.35), p=0.735, ES = 0.07), and for patients with other diagnoses (N=28), it had decreased by 3.39 points (SD=9.74), p=0.0763, ES = -0.35). Of the 26 patients who completed the PGIC, seven reported that their condition worsened, 10 perceived no change, and nine reported an improvement of their condition.

## Discussion

The results of this study show that key outcomes experienced in daily life can be reported by patients at early or intermediate stage of a cSVD using a simple questionnaire limited to fewer than 20 questions. This instrument was developed in individuals with ischemic cSVD, either sporadic or genetic, including CADASIL patients. The results demonstrate that the final tool was particularly fitted for CADASIL patients; hence, we chose the acronym CADA-PRO (CADASIL patient reported outcome) to name this questionnaire. To our knowledge, this is the first instrument that can help measure, in a comprehensive manner, the cognitive, motor, emotional and behavioral impact of a cSVD on the quality of life, as perceived by the patients themselves. Different PRO measures have been previously developed for stroke patients. Some of these tools, as the Stroke and Aphasia Quality of Life Scale-39 item version (31), were strongly focused on a specific deficit encountered after large cerebrovascular lesions not encountered in cSVD patients. Others such as the Stroke Satisfaction Care (32) were prepared for evaluating specific services, such as rehabilitation, after persisting stroke deficits. Various multidimensional tools, such as the Newcastle Stroke-Specific Quality of Life Measure (33), were found unrelated to the family support or treatment effect. Finally, even the Stroke-PROM (34), one of the most comprehensive measures, was validated only in elderly individuals and included questions related to physical deficits that are not relevant to cSVD. Thus, none of the existing tools designed for cerebrovascular diseases was specifically built on the complaints of patients with cSVD, as proposed herein.

In contrast to many PRO tools whose quality metrics have rarely been extensively evaluated, the CADA-PRO questionnaire has been developed not only rigorously but also following a multistep procedure and the latest COSMIN guideline recommendations (16). In the present study, the administration of the final 18-items CADA-PRO tool showed few missing data, no ceiling or floor effect, and an excellent internal consistency. Through a deep analysis of the large data collected, we identified four main dimensions with significant impact from the patient’s point of view, corresponding to the 4-domain structure of the scale, assessing 1 - disturbances in common activities of daily life, 2 - difficulties related to attention and executive function deficit, 3 - motor symptoms including focal deficits or gait disturbances, and finally 4 - anxiety and depression. These features correspond well to frequent complaints of patients at early or intermediate stage of the disease. Most items of the CADA-PRO tool were focused on executive dysfunction, mood and behavior, rather than late symptoms such as dementia or motor dependency, as they were developed for and by individuals recruited in consultations or short-term hospitalization, who were able to complete the questionnaires themselves. The most seriously ill patients, who were bedridden or unable to go to the Referral Centre, could not be included in such a study. Consequently, the CADA-PRO should not be considered as a tool that summarizes all potential consequences of a cSVD, but rather as one that captures the main difficulties experienced by the patient before the latest disease stage. As anosognosia is expected to appear very late in the course of a cSVD, the choice of focusing on patients at early or intermediate disease stage to develop our tool also proved reasonable. Moreover, the subjects recruited in the study were clearly the most relevant target for future testing of preventive therapeutic interventions long before the occurrence of dependency. Furthermore, the CADA-PRO questionnaire presents a good test-retest reliability, but responses to some items could vary when the questionnaire was administered at hospital or one month later at home. These findings suggest that the environment context, the family closeness, or perhaps the distance from the care team or structure might also influence some responses. In practice, the CADA-PRO instrument should be administered in a constant environment for obtaining the best reliability. Finally, no significant change in patients’ perceived condition between inclusion and the one-year follow-up was observed neither using CADA-PRO score nor the PGCI. These results indicate that the patients included in the study did not experience a net worsening of their disease over one year, in line with the relatively slow progression of the disease over decades. Thus, a larger sample size and longer follow-up would be required to answer the key question of how patients perceive the progression of their disease.

An important finding of our study concerns the contrasting results obtained in CADASIL patients and in those having a different cSVD. In the group of subjects with confirmed NOTCH3 mutations, the CADA-PRO total score was significantly correlated with the Rankin score, the SAS Apathy scale, HAD scores as well as TMT times. The Anxiety-Depression sub-score was significantly correlated with the HAD and SAS scores, reflecting mood alterations and apathy respectively. In the same group of patients, sub-scores from the other domains were also significantly correlated with the Working Memory Index from WMS-III or with TMT A time. These correlations were not observed in patients with cSVDs distinct from CADASIL. In this group, the CADA-PRO score and its sub-scores were poorly correlated with cognitive tests or motor disability as assessed using the modified Rankin score. The total score was significantly correlated only with the HAD and SAS scores. Altogether, these results indicate that the CADA-PRO tool is well fitted for CADASIL patients to report key outcomes in strong agreement with multiple facets of bedside clinical evaluation. Patients with other cSVDs furthermore showed some differences from patients with CADASIL: older age at inclusion and diagnosis, worse performances in several neuropsychological assessments. Additional questionnaire items, larger samples or further investigations are likely needed for improving the PRO measure for other types of cSVD.

Finally, we found that the total CADA-PRO score was only weakly correlated with the different cognitive scores. These results are consistent with the lack of association between the subjective cognitive complaints and neuropsychological performances reported in 152 patients with white matter hyperintensities (35). They are also in agreement with the results of a meta-analysis showing only a weak link between cognitive complaints and cognitive scores in the elderly (36), and an independent effect of depressive symptoms on cognitive performances (35, 36). Hence, the CADA-PRO questionnaire may not reflect the extent of cognitive difficulties associated with the disease. In CADASIL patients, the association of the CADA-PRO total score with the working memory index was independent of the association with the HAD depression score. Therefore, the emotional and behavioral impact of the disease appears to influence the patients’ quality of life in ways that are distinct from cognitive impairment, at least as measured using the CADA-PRO tool. These emotional and behavioral symptoms should not be overlooked, as in patients with Cerebral Small Vessel Disease, mood disturbances were shown to be the first predictor of quality of life (37).

This study comprises several methodological strengths. The CADA-PRO questionnaire was developed from real data, in close association with cSVD or CADASIL patients and their families, and with experts having a long experience in the management of genetic or sporadic cSVD. Two samples of patients were recruited, with an acceptable number of individuals affected by a rare disease. Strict criteria were used for questionnaire item selection and a wide range of psychometric properties were thoroughly investigated according to the best and updated recommendations for the development of PRO measures. The present study also suffers from a number of limitations. The sample of individuals with a cSVD distinct from CADASIL was relatively limited. This might prevent obtaining a PROs instrument enabling to cover all potential phenotypic aspects of multiple sporadic or genetic cSVDs. Moreover, the CADA-PRO instrument could not be used by both the patient and his/her caregivers. To account for the absence of a caregiver in 70% of our patients, further development of a caregiver-reported outcome using a larger sample would be promising. Our development study also showed that disagreement between patient and caregiver was significantly higher when the average CADA-PRO score increased, suggesting that anosognosia may develop in some patients as the disease progresses. This is consistent with previous results showing that informant-reported cognitive complaints, but not patient-reported cognitive complaints, correlate with white matter hyperintensities volume and functional abilities in cSVD patients (35). Furthermore, our tool would benefit from assessing its correlation with another scale measuring quality of life. A last limitation is that the CADA-PRO questionnaire was developed in France and should now be translated and further validated for their use in multiple countries.

In summary, the CADA-PRO tool can now be used in CADASIL patients to assess patient-perceived impact of their disease in cross-sectional and longitudinal studies. Measures derived from this tool have been deeply analyzed. This instrument is largely validated and appears now suitable to provide additional secondary endpoints for future clinical trials. Currently, the CADA-PRO tool is only available in the French language and should now be urgently translated to allow its use worldwide. Additional efforts are also needed to develop a caregiver version of PROs in CADASIL from the present instrument.

## Non-standard Abbreviations and Acronyms

CADASIL: cerebral autosomal dominant arteriopathy with subcortical infarcts and leukoencephalopathy
cSVD: cerebral Small Vessel Disease
EF: Executive Functions
EFA: Exploratory Factor Analysis
FCSRT: Free and Cued Selective Reminding Test
HAD: Hospital Anxiety and Depression scale
MIRT: Confirmatory Multidimensional Item Response Theory
mRS: modified Rankin scale
PGIC: Patient Global Impression of Change
SAS: Starkstein Apathy Scale
SSPB - 4: Short Physical Performance Battery (4 items)
TMT: Trail Making Test
VADAS-Cog: Vascular Dementia Assessment Scale cognitive subscale
SDMT: Symbol-Digit Modalities Test
WCST: Wisconsin Card Sorting Test
WMS-III: Weschler Memory Scale 3^rd^ revision

## Acknowledgment

We thank the team in charge of formatting and cleaning the database Mrs Claire Pacheco; Mr Abbas Taleb for managing and collecting the data; Mrs Solange Hello and Celine Martin who managed and organised the appointment of multiple family members involved in the present study; Mrs Nathalie Gastelier, the research manager in charge of the Cohort Study. We also thank the CADASIL France Association for their help and permanent support.

## Supplementary Materials

Tables S1–S4

Figure S1–S4

